# Development of a Clinical Speech Corpus with Temporally Aligned Sensitive Health Information Annotations

**DOI:** 10.64898/2026.03.31.26349906

**Authors:** Hsieh-Chih Hsu, Liang-Chun Fang, Tatheer Hussain Mir, Ching-Tai Chen, Hui-Hsien Feng, Jiunn-Ru Lai, Pratham Nandy, Omkar Panchal, Wei-Hsiang Liao, You-Zhen Tien, Pin-Zhen Chen, Yun-Ru Lin, Hong-Jie Dai, Jitendra Jonnagaddala

## Abstract

**Objectives:** Publicly available datasets for clinical speech de-identification remain scarce due to privacy constraints and annotation complexity. We address this gap with the 2025 SREDH/AI Cup SHI Speech Corpus, a time-aligned clinical speech dataset for detecting sensitive health information (SHI).

**Methods:** The dataset integrates three sources: the OpenDeID-v2 corpus, the dataset for automated medical transcription and medical dialogues derived from Taiwanese television dramas, combining English medical speech with Mandarin Chinese utterances. All audio was manually annotated with millisecond-level, time-aligned SHI segments using a schema of eight primary categories and 38 subcategories of which 26 were attested.

**Results:** The corpus comprises approximately 20 hours of annotated audio (19.36 hours English, 0.89 hours Chinese) and 7830 SHI entities, split into training (10 hours, 1539 files), validation (5 hours, 775 files) and test (5 hours, 710 files) sets, supporting evaluation of both automatic speech recognition (ASR) and SHI recognition. Interannotator agreement (Fleiss’s kappa) reached 0.907. In the 2025 SREDH/AI Cup competition, top-performing systems achieved a mixed error rate of 0.1147 in ASR and a macro F1 of 0.7103 in SHI recognition.

**Discussion:** SHI category distribution is long-tailed, and English–Chinese data volume is highly imbalanced, underscoring the scarcity of Chinese clinical speech resources and the difficulty of time-aligned annotation in low-resource settings.

**Conclusion:** This corpus approximates dialogue-based clinical scenarios and supports research on medical speech de-identification, offering a unique reference for low-resource clinical speech privacy protection.

**Key message:** *What is already known on this topic:*

- Publicly available clinical speech datasets containing time-aligned sensitive health information (SHI) annotations for clinical speech de-identification research are limited.

*What this study adds:*

- The 2025 SREDH AI Cup SHI Speech Corpus introduces a clinically oriented speech dataset with millisecond-level time-aligned SHI annotations covering 26 of the 38 SHI subcategories defined in the annotation schema.
- The corpus integrates structured annotation protocols and standardized data processing to support the reproducible benchmarking of speech-based de-identification models.

*How this study might affect research, practice or policy:*

- The availability of time-aligned SHI annotations may facilitate research on real-time or streaming de-identification systems beyond conventional, transcription-focused approaches.
- This dataset may support the development of privacy-preserving technologies for clinical speech in bilingual settings involving English and low-resource languages.

## INTRODUCTION

Clinical data contains a wealth of information valuable for research,^1,2^ and to prevent the leakage of private information, sensitive health information (SHI) must be removed or replaced with the corresponding surrogates before such data can be used. The 2014 i2b2^3^ clinical de-identification natural language processing challenge released a corpus of 1304 medical records from 296 patients with diabetes. These records contain detailed clinical narratives annotated for SHI. The MIMIC-III de-identification dataset ^4^ comprises 60 725 manually annotated SHI entities, categorised into six major classes and 22 specific types. In addition, the OpenDeID-v2 ^5,6^ corpus, released during the 2023 SREDH/AI Cup competition, was constructed from 3244 electronic medical records and includes annotations for SHI and temporal normalisation. Although these resources are well suited for training a named entity recognition (NER) model for the de-identification task, they are text-based and therefore cannot be directly applied to speech de-identification.

Existing speech corpora like LibriSpeech,^7^ Common Voice,^8^ AISHELL-1^9^ and MagicData-RAMC^10^ are predominantly compiled for automatic speech recognition (ASR) rather than de-identification. SLURP^11^ is a 58-hour English speech dataset spanning 18 domains, 46 intents and 54 entity types, including person and datetime information. AISHELL-NER^12^ extends the AISHELL-1 corpus by annotating three common entity types: names, organisations and locations. Nevertheless, these datasets are primarily designed for general-domain spoken language understanding, and their linguistic coverage does not align well with the characteristics of clinical communication. More importantly, their annotations are entity-centric rather than temporally grounded, lacking information about where each entity occurs in the speech signal. Therefore, a key gap in existing research is the lack of timestamp-level entity annotations for clinical speech.^12^

To address this gap, we compiled the 2025 Secure Research Environment for Digital Health (SREDH)/AI Cup SHI Speech Corpus as a domain-specific evaluation resource for speech de-identification. This corpus is primarily designed as an English clinical speech benchmark, with a smaller Mandarin Chinese subset as a supplementary bilingual extension. Its content simulates dialogue-based clinical scenarios, and all recordings are accompanied by human-verified transcripts and time-aligned annotations covering 26 of the 38 SHI subcategories defined in the annotation schema. This design supports English-centric evaluation while providing an initial basis for future research on bilingual and code-mixed clinical speech de-identification.

## METHODS

### Data source

The 2025 SREDH/AI Cup SHI Speech Corpus was constructed by integrating and extending complementary medical-domain resources from the OpenDeID-v2 corpus,^6^ the dataset for automated medical transcription (DAMT)^13^ and clinical-style dialogue materials from licensed videos from Taiwanese television dramas provided by Taiwan Public Television Services (PTS).

The OpenDeID-v2 corpus was originally released for the 2023 SREDH/AI Cup^6^.It comprised unstructured electronic medical record (EMR) reports derived from Australian hospitals^14^. While this corpus provides comprehensive text-level SHI annotations, it lacks corresponding speech recordings. In contrast, DAMT contains high-quality psychiatric dialogue recordings generated from scripted clinical scenarios. However, it lacks explicit SHI entity annotations necessary for structured de-identification modelling.

Given that both datasets were English-only, we incorporated clinical-style Chinese-oriented dialogue audios from PTS to enhance linguistic diversity and real-world applicability. The resulting bilingual dataset, which combines English-dominant resources with Mandarin Chinese data, better reflects authentic clinical communication patterns observed in Taiwanese healthcare contexts. By integrating these three data sources, we established a unified dataset containing audio recordings with temporally aligned SHI annotations.

### Corpus development

Table 1 Presents the annotation schema defined for compiling the 2025 SREDH/AI Cup SHI Speech Corpus. The schema is primarily based on our previous guidelines^6^, comprising eight primary categories and 38 subcategories, of which 26 are attested in the corpus (table 3). Online supplemental table S1 illustrates the annotation of an actual corpus segment, showing, for each audio file, the SHI type, the start and end timestamps and the corresponding surface form of each annotated SHI entity. All entries in Table 1 and online supplemental table S1 are replaced by surrogates^.15^ None of them were derived from any real individual’s records or experiences. In addition, the examples in table 1 are independent illustrations of individual SHI categories and do not correspond to any single individual; the profession, medical record number and other fields are not attributes of the same person. Accordingly, no entry in table 1 can be linked back to, combined into a profile of or used to reidentify any real person.

**Table 1.** Health Science Alliance SHI annotation schema.

| SHI category | Subcategory | Example |
| --- | --- | --- |
| NAME | PATIENT, DOCTOR, USERNAME, PERSONALNAME, AND FAMILYNAME | Gudara, Pineau,<br>Shaunda, and Hinck |
| PROFESSION | (none) | Lawyer and teacher |
| LOCATION | ROOM,DEPARTMENT,HOSPITAL,ORGANIZATION, STREET, CITY, DISTRICT, COUNTY, STATE, COUNTRY, ZIP, OTHER | PERIOPERATIVE UNIT-POW, MACQUARIE<br>WARD - RHW |
| AGE | (none) | 23, 98 |
| DATE | DATE, TIME, DURATION, AND SET | September 26 |
| CONTACT | PHONE, FAX, EMAIL, URL, AND | , |
|  | IP ADDRESS | 194.223.1.1 |
| IDs | SOCIAL_SECURITY_NUMBER, MEDICAL_RECORD_NUMBER, HEALTH_PLAN_NUMBER, ACCOUNT_NUMBER, LICENSE_NUMBER, VEHICLE_ID,DEVICE_ID, BIOMETRIC_ID, AND ID_NUMBER | MRN: 9174338 |
|  |  | IDNUMBER:<br>**12R1500257** |
| OTHER | (none) | Fingerprint, Photograph, and Company Logo |

Due to the heterogeneity in data formats and file structures across the three data sources, each underwent source-specific preprocessing before being annotated by the five trained annotators (L-CF, W-HL, Y-ZT, P-ZC and Y-RL) for time-aligned SHI spans.

The DAMT already contains speech recordings paired with corresponding transcriptions; therefore, no additional data selection or filtering was applied to this subset. For the PTS-derived audio, we manually screened the recordings and retained only the dialogue segments depicting clinical or medical scenarios. Segments containing non-clinical content, excessive background audio or unrelated scenes were excluded prior to annotation. The retained speech segments were subsequently transcribed and annotated with SHI.

On the other hand, the OpenDeID-v2 corpus consists exclusively of text-based EMR reports annotated with surrogate SHIs^6 15^. A key advantage of leveraging this resource was that all SHI categories had already been professionally annotated and replaced with surrogate values, eliminating the need for additional SHI reannotation while preserving privacy safeguards. However, as the corpus does not include corresponding speech recordings, the text reports were converted into spoken-language scripts to enable natural speech-based modelling. A subset of 300 EMR reports was randomly sampled and rephrased into recording scripts by two domain specialists. During this process, the original surrogated SHI annotations were strictly preserved to maintain annotation consistency. Each script, approximately 100–200 words in length, was adapted to simulate natural clinical discourse while retaining the original entity structure.

The prepared scripts were recorded by 25 participants (9 males and 16 females), each contributing 10–20 audio samples. To ensure interspeaker consistency, all participants rehearsed their assigned scripts independently and received standardised instructions regarding speech rate, pause placement and pronunciation. The resulting recordings, ranging from 1.0 to 1.8 min, were maintained and underwent rigorous quality control before downstream processing. Data acquisition took place in quiet, enclosed environments, with all samples captured using smartphone devices.

After preparing transcripts and audio from all sources, a unified processing pipeline was implemented. Forced alignment synchronised text and audio based on the Montreal forced aligner,^16^ using the English (USA) Advanced Research Projects Agency (ARPA) acoustic and dictionary models (V.3.0.0) for English data, and the Mandarin Montreal Forced Aligner (MFA) acoustic and dictionary models (V.3.0.0) for Mandarin data. Subsequently, voice activity detection was applied to segment recordings into clips under 30 s. Each segment was then aligned with its corresponding SHI text spans. Figure 1 illustrates the complete pipeline of this process, spanning from data collection and preprocessing through forced alignment to final data annotation across the three data sources. The annotation process was supported by Label Studio^17^ as illustrated in figure 2. SHI entities were identified based on auditory inspection according to the annotated texts supported by waveform visualisations to determine precise temporal boundaries. Start and end timestamps were recorded at millisecond resolution.

**Figure 1.**
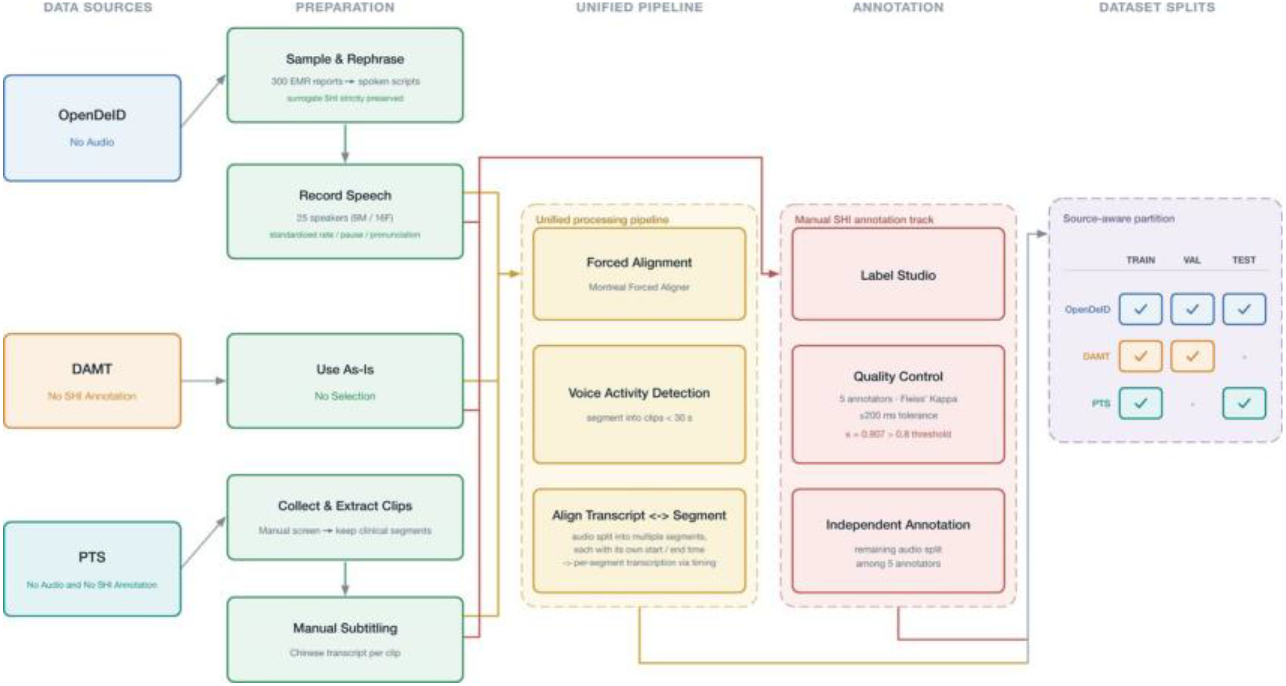
The creation pipeline of the 2025 SREDH/AI Cup SHI speech corpus. DAMT, dataset for automated medical transcription; EMR, Electronic Medical record;PTS, Public Television Services; SHI, sensitive health information; SREDH, Secure Research Environment for Digital Health.

**Figure 2.**
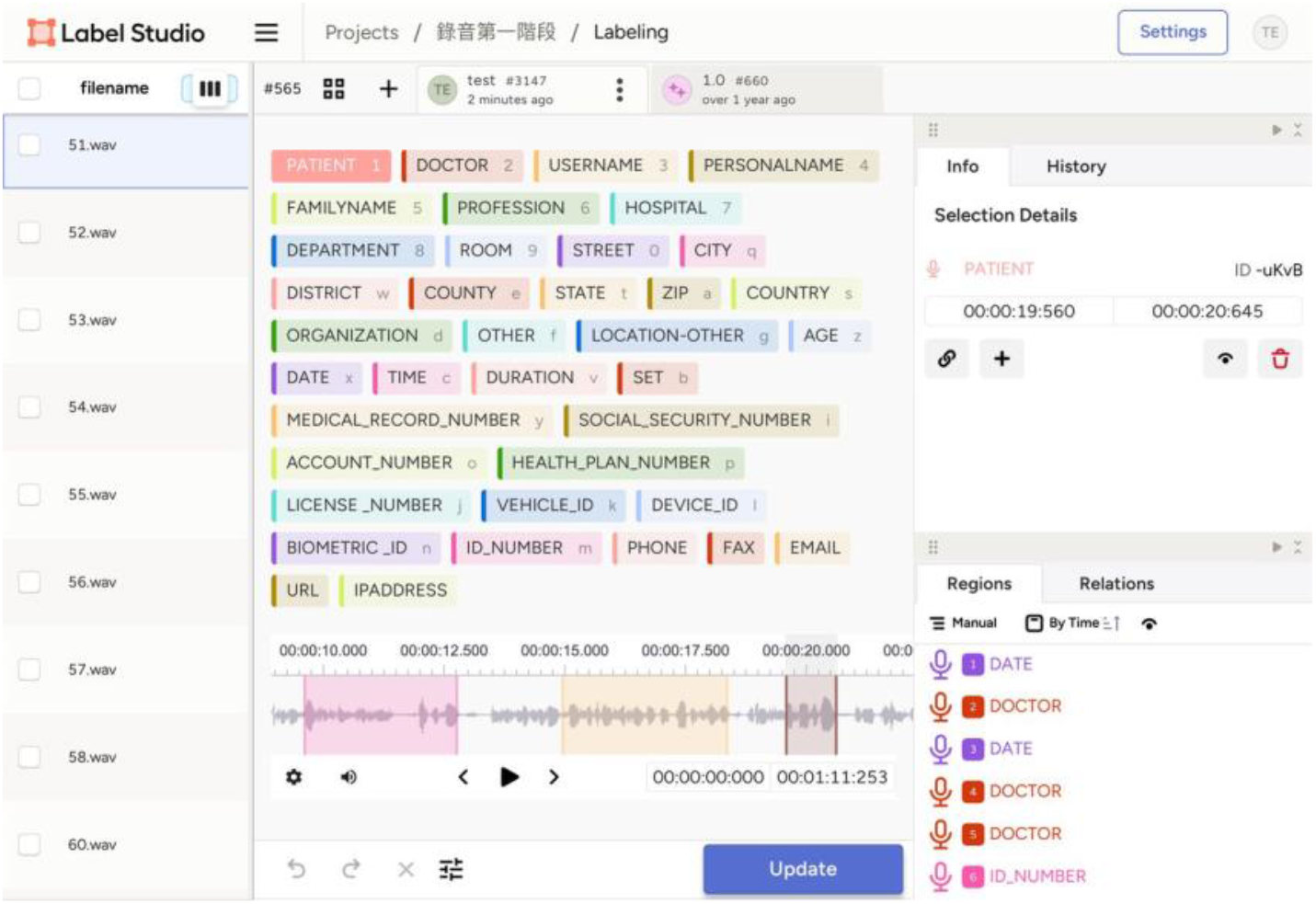
Label Studio interface used for SHI annotation, showing synchronised audio playback, waveform visualisation, corresponding transcription and time-aligned SHI span. SHI, sensitive health information.

To account for the inherent variability in manual temporal annotation performed by five annotators, agreement was measured using Fleiss’s Kappa. Following the evaluation principles established in the NIST Rich Transcription 2006 guidelines,^18^ which recommend temporal tolerance to accommodate minor boundary discrepancies, a ±200 ms tolerance window was applied when determining boundary agreement. Two annotations were considered matching if both their start and end timestamps fell within this tolerance range.

The annotation process was conducted in two phases. During the training phase, approximately 8 hours of speech from 20 DAMT reports was jointly annotated by all five annotators. After 12 iterative calibration rounds, a Kappa value of 0.907 was achieved, exceeding the predefined acceptance threshold of 0.8. Subsequently, the remaining data were evenly distributed among the annotators for formal annotation.

For the 2025 SREDH/AI Cup competition, the compiled corpus was partitioned into training, validation and test sets according to the data source and intended evaluation scenario. For the PTS-derived Chinese audio, the selected clinical dialogue segments were randomly split by duration into training and test sets using a 1:1 ratio, respectively. The split was performed at the segment level after non-clinical content, background audio and unrelated scenes were excluded.

For the English data, the test set was designed to reflect the target application scenario of speech de-identification from clinical-style spoken content. Accordingly, the English test set consisted only of self-recorded audio generated from rephrased OpenDeID-v2 corpus reports, whereas DAMT recordings were excluded from the English test set and used only for training and validation. This source-aware design reduced potential overlap between training and test scripts and mitigated the risk of information leakage, given that DAMT is publicly available and its reference transcriptions could potentially be accessed by participants in advance. The resulting partition enabled separate evaluation of transcription performance and temporally aligned SHI recognition under a setting closer to the intended use case.

## RESULTS

The 2025 SREDH/AI Cup SHI Speech Corpus comprises approximately 20 hours of audio (20.25 hours in total), including DAMT (59%), OpenDeID-v2 corpus (36%) and PTS-based scripted clinical dialogues (5%). This composition yields 19.36 hours of English and 0.89 hours of Chinese speech. Please note all percentages are rounded to the nearest integer and durations to two decimal places; rounded figures may therefore not sum exactly to the stated totals. The corpus was partitioned into training, validation and test sets of approximately 10, 5 and 5 hours, respectively. The English corpora used in this study exhibit distinct accent characteristics based on their sources. While the DAMT dataset lacks explicit linguistic labelling, it is presumed to predominantly feature American English; conversely, the English audio in OpenDeID-v2 corpus primarily consists of Taiwanese English accents. Regarding the Chinese corpora, all recordings consistently feature Taiwanese Mandarin. All subsets maintained a signal-to-noise ratio (SNR) above 28 dB, indicating sufficient acoustic clarity for downstream processing. Speech rate across the three data sources was quantified using syllables per second (SPS). Statistical analysis revealed that the mean SPS for all three sources consistently ranged between 3.233 and 3.867, indicating a highly comparable speaking pace across the datasets. Furthermore, univariate analysis of the SPS metric demonstrated distinct patterns in data dispersion: the SD of SPS for both English sources was close to 1.0, aligning with a standard distribution. In contrast, the Chinese dataset exhibited a substantially larger SD, a finding that reflects the pronounced variability introduced by collecting Chinese speech samples from diverse dramatic sources. Detailed file counts and SNR statistics for each subset are provided in table 2.

**Table 2.** Detailed information for the ASR task in the 2025 SREDH AI Cup SHI Speech Corpus.

| Subset | Duration | Number of files | Mean SNR | Median SNR | Std SNR | Min SNR | Max SNR |
| --- | --- | --- | --- | --- | --- | --- | --- |
| Training | 10 hr | 1,539 | 42.67 dB | 42.35 dB | 12.85 dB | 3.71 dB | 77.51 dB |
| Validation | 5 hr | 775 | 39.14 dB | 37.12 dB | 14.23 dB | 12.70 dB | 90.20 dB |
| Test | 5 hr | 710 | 28.71 dB | 28.78 dB | 6.73 dB | 6.55 dB | 52.41 dB |

| Language | Duration | Number of files |
| --- | --- | --- |
| English | 19.36 hr | 2,875 |
| Chinese | 0.89 hr | 149 |

| Data Source | Language | Mean SPS | Std SPS | Number of files | Duration | Contribution (%) |
| --- | --- | --- | --- | --- | --- | --- |
| The OpenDeID v2 corpus | English | 3.233 | 0.573 | 1,095 | 7.36 hr | 36 |
| DAMT | English | 3.867 | 0.725 | 1,780 | 12.00 hr | 59 |
| PTS | Chinese | 3.606 | 2.975 | 149 | 0.89 hr | 5 |

Given the fine-grained and temporally localised nature of SHI annotation in audio, maintaining annotation consistency was essential to ensure dataset reliability. After 12 iterative calibration rounds, the annotation team achieved a Fleiss’s Kappa of 0.907. Following these protocols, 7830 SHI entities across 26 subcategories were annotated across the 20-hour corpus for the SHI recognition task. Several entity categories occurred at very low frequencies, resulting in a pronounced long-tail distribution, as illustrated in table 3.

**Table 3.** Distributions of 26 SHI subcategories in the 2025 SREDH AI Cup SHI Speech Corpus.

|  | Training set | Validation set | Test set | Total |
| --- | --- | --- | --- | --- |
| PATIENT | 178 | 182 | 468 | 828 |
| DOCTOR | 227 | 306 | 832 | 1,365 |
| PERSONALNAME | 194 | 46 | 4 | 244 |
| FAMILYNAME | 146 | 28 | 12 | 186 |
| PROFESSION | 26 | 16 | 2 | 44 |
| HOSPITAL | 40 | 73 | 167 | 280 |
| DEPARTMENT | 39 | 67 | 135 | 241 |
| ROOM | 3 | 0 | 2 | 5 |
| STREET | 24 | 64 | 116 | 204 |
| CITY | 55 | 65 | 120 | 240 |
| **DISTRICT** | 0 | 1 | 0 | 1 |
| **COUNTY** | 2 | 2 | 0 | 4 |
| **STATE** | 40 | 64 | 111 | 215 |
| **ZIP** | 22 | 60 | 115 | 197 |
| **COUNTRY** | 11 | 3 | 2 | 16 |
| **ORGANIZATION** | 20 | 12 | 6 | 38 |
| **LOCATION-OTHER** | 11 | 6 | 4 | 21 |
| **AGE** | 40 | 17 | 34 | 91 |
| **DATE** | 668 | 493 | 650 | 1,811 |
| **TIME** | 171 | 110 | 125 | 406 |
| **DURATION** | 336 | 152 | 8 | 496 |
| **SET** | 41 | 29 | 8 | 78 |
| **MEDICAL_RECORD_NUMBER** | 31 | 68 | 158 | 257 |
| **ID_NUMBER** | 60 | 175 | 324 | 559 |
| **PHONE** | 1 | 0 | 1 | 2 |
| **URL** | 1 | 0 | 0 | 1 |

This corpus was adopted as the benchmark dataset for the 2025 SREDH/AI Cup Medical Speech Sensitive Information Recognition Competition,^19–21^ which aimed to evaluate state-of-the-art systems for ASR and SHI recognition in clinical speech. The challenge was organised into two sequentially linked subtasks, each targeting a distinct capability required for speech-based de-identification. The ASR subtask required systems to produce faithful transcriptions of clinical dialogues, whereas the SHI recognition subtask required systems to detect, temporally localise and classify SHI mentions from the spoken input. The top-performing team^22^ adopted a dual-modality pipeline approach. For the ASR subtask, they fully fine-tuned CrisperWhisper^23^ and FireRedASR^24^ as the English and Chinese ASR models respectively. For the SHI recognition subtask, they used Qwen3^25^ as the backbone and applied NEFTune,^26^ which injects random noise into the input embeddings during fine-tuning to improve generalisation. Online supplemental table S2 summarises the final results of the top five teams. Whisper was the most widely adopted model for the ASR subtask, whereas the SHI recognition methods diverged into two contrasting paradigms: generative approaches, represented by Qwen 3 and Llama 3, and semantic-understanding approaches based on BERT-style encoders such as DeBERTa and RoBERTa. Across the benchmark, the rank-2 team achieved the best mixed error rate (MER) of 0.1147 in the ASR subtask, while the rank-1 team achieved the highest macro F1-score of 0.7103 in the SHI recognition subtask. In contrast, the average performance across participating systems was considerably lower, with an MER of 0.3539 and a macro F1 of 0.2696. The discrepancy between leading and average systems underscores the complexity of the task, especially when moving beyond transcription toward temporally aligned recognition of clinically sensitive entities. These results demonstrate that the 2025 SREDH/AI Cup SHI Speech Corpus serves not only as a shared evaluation resource but also as a challenging benchmark for developing robust, clinically grounded and clinical speech de-identification methods under the English and low-resource bilingual conditions.

## DISCUSSION

The 2025 SREDH/AI Cup SHI Speech Corpus was developed to address two important gaps in existing resources: the limited availability of speech data for deidentification research and the lack of temporally aligned SHI annotations in clinical speech datasets. Because clinical speech data suitable for de-identification research remains critically scarce, we constructed a controlled clinical-style speech corpus by reformulating OpenDeID-v2 corpus reports into spoken-language scripts, incorporating existing clinical-style dialogue recordings and adding selected Chinese dialogue materials. The resulting corpus provides audio recordings with human-verified transcripts and time-aligned SHI annotations, thereby supporting the development and evaluation of speech-based de-identification methods.

Certain categories of SHI appear far less frequently than others, forming a long-tail pattern. This pattern is not incidental; rather, it reflects both the properties of the source materials and the distributional characteristics commonly observed in clinical documentation. For example, CONTACT entities appear sparsely, consistent with the previous study,^14^ which noted that clinical records carry limited patient-related attributes such as occupation or contact information. Based on our observations, the DAMT data contains almost no ID-related entities. Prior work^27^ has similarly shown that publicly available de-identification datasets tend to overrepresent common SHIs such as names, while underrepresenting less frequent categories such as passport numbers and identification numbers. This imbalance has important implications for model development and evaluation, as performance on common SHI categories may not fully reflect a system’s ability to recognise rare but privacy-critical entities.

Beyond the distribution of SHI categories, the corpus is also shaped by its linguistic composition. The corpus was developed primarily as an English-dominant evaluation resource, with Chinese data included as an extension motivated by the prevalence of bilingual communication in certain clinical settings.^28^ Despite this motivation, the dataset is heavily skewed toward English, reflecting the broader scarcity of annotated Chinese clinical speech resources. The compilation process itself illustrates this scarcity. Although audio from Chinese films and dramas is abundant, most candidate materials were not suitable for inclusion. As Tannen observes in her book Talking Voices,^29^ dramatic dialogue is a distilled and refined version of reality rather than a direct imitation of everyday interaction, and personal identifiers are often deliberately omitted to preserve narrative flow. After applying SHI-compliance criteria, only 1 hour of Chinese data could be retained. This limited yield underscores both the shortage of usable Chinese medical speech resources and the value of the Chinese subset included in this corpus.

Given the size and composition of the Chinese subset, it should be understood as a complement to the English-dominant corpus rather than as a parallel monolingual benchmark. The corpus is therefore better suited for studying bilingual or code-mixed speech de-identification under a unified processing framework than for drawing strong conclusions about language-specific performance in Chinese alone. With multilingual foundation models such as Whisper^30^ and Qwen3-ASR^31^ increasingly capable of processing multiple languages within a single pipeline, the corpus may support future studies that jointly examine ASR and SHI recognition performance. However, quantitative comparisons between the unified bilingual system and language-specific alternatives are beyond the scope of the present corpus construction study and should be explored in future work.

Finally, although our controlled corpus construction design offers practical advantages for data sharing while reducing the ethical and privacy risks associated with releasing real patient–clinician conversations, it should be noted that the corpus was not collected from naturally occurring clinical encounters. Instead, it was constructed from rephrased clinical-style scripts, controlled re-recordings and drama-derived clinical-style dialogues. As a result, it may not fully capture the spontaneity, acoustic variability, interruptions, overlapping speech, emotional dynamics and interactional complexity commonly observed in real clinical conversations. The corpus is therefore best understood as a controlled clinical-style speech resource for developing and evaluating speech de-identification methods, rather than as a fully representative sample of real-world clinical communication. Accordingly, findings derived from this corpus should not be directly extrapolated to real-world clinical deployment. Future work should incorporate naturally occurring clinical recordings, where feasible and ethically permissible, to further improve ecological validity and generalisability. We make no claims regarding pooled multilingual evaluation, cross-lingual transfer or real-world deployment performance; these settings remain important directions for future investigation.

## Data Availability

Researchers wishing to use the corpus should refer to https://github.com/SREDH-Consortium/2025-SREDH-AICup-SHI-Speech-Corpus for data access and follow the instructions provided on the page to apply for a data use license. For OpenDeID v2 Corpus,see - https://github.com/SREDH-Consortium/OpenDeID-Corpus For the Chinese-language data, as the drama content is under the copyright of the respective television networks, we can only provide the annotation files along with index files indicating the corresponding locations in the original videos.

## Acknowledgments

We would like to express our gratitude to the University of New South Wales, the Lowy Cancer Research Center, and the HSA Biobank for granting access to the medical records used in this study. We acknowledge the IW-DMRN workshop (https://www.sredhconsortium.org/sredh-workshops/2025-iw-dmrn/submission-information) and the SREDH Consortium(https://www.sredhconsortium.org/), particularly the Translational Cancer Bioinformatics working group, for providing access to the OpenDeID corpus v2 dataset and for their valuable contributions to this research. We also thank PTS for granting permission to use audio excerpts from its licensed drama content.

## Contributors

Contributors: H-JD, C-TC, H-HF, J-RL and H-CH designed the study. H-CH, L-CF and THM made substantial contributions to the summary and analysis of the survey information. L-CF, W-HL, Y-ZT, P-ZC and Y-RL made substantial contributions to data annotation. PN and OP provided critical comments and revisions to the manuscript. H-CH drafted the initial manuscript. H-JD and JJ supervised the study and critically revised the manuscript. JJ acted as guarantor and takes responsibility for the integrity of the work. All authors contributed to the revision of the manuscript, approved the final version for submission and agree to be accountable for all aspects of the work.

## Ethics approval

UNSW Sydney Human Research Ethics Committee Approval No. HC17749.

## Data availability statement

Researchers wishing to use the corpus should refer to https://github.com/SREDH-Consortium/2025 SREDH AI Cup SHI Speech Corpus for data access and follow the instructions provided on the page to apply for a data use license. For OpenDeID v2 corpus,see - https://github.com/SREDH-Consortium/OpenDeID-Corpus For the Mandarin Chinese data, as the drama content is under the copyright of the respective television networks, we can only provide the annotation files along with index files indicating the corresponding locations in the original videos.

